# Long-term monitoring of SARS-CoV-2 load and variant composition at a large metropolitan wastewater treatment plant using a simple two-step direct capture RNA extraction, droplet digital PCR, and targeted mutation assays

**DOI:** 10.1101/2024.08.21.24311866

**Authors:** Steven J. Balogh, George B. Sprouse, Kenneth B. Beckman, Ray H.B. Watson, Darrell M. Johnson, Lee D. Pinkerton, Yabing H. Nollet, Adam W. Sealock, Walter S.C. Atkins, Laura M. Selenke, Joseph A. Kinney, Patrick J.R. Grady, Brandon Vanderbush, Jerry J. Daniel

**Affiliations:** Metropolitan Council Environmental Services, 2400 Childs Road, Saint Paul, MN, 55106, USA; University of Minnesota Genomics Center, Minneapolis, MN, 55455, USA

**Keywords:** COVID-19, wastewater-based epidemiology, virus, molecular analysis, Minnesota

## Abstract

Wastewater surveillance offers an objective, comprehensive, and cost-effective means of monitoring the prevalence and genomic heterogeneity of pathogens circulating in a community. Here, a novel two-step extraction procedure for the direct capture of SARS-CoV-2 RNA from raw wastewater is presented. Combined with reverse transcription-droplet digital polymerase chain reaction (RT-ddPCR) detection, the method provides a fast and sensitive method for measuring viral RNA concentrations in wastewater. The method was used to measure the concentration of SARS-CoV-2 RNA in daily samples of wastewater entering a major metropolitan wastewater treatment plant over the course of 32 months, from November 2020 through June 2023. In addition, targeted mutation assays were used with RT-ddPCR to characterize the evolving presence and prevalence of specific SARS-CoV-2 variant sub-lineages in the wastewater stream over time. The results demonstrate the utility of these methods to accurately measure the total load of SARS-CoV-2 RNA, and chronicle its evolving variant composition, in wastewater treatment plant influent, providing near-real-time characterization of COVID-19 disease prevalence and trends in the served community.

**Graphical abstract:** 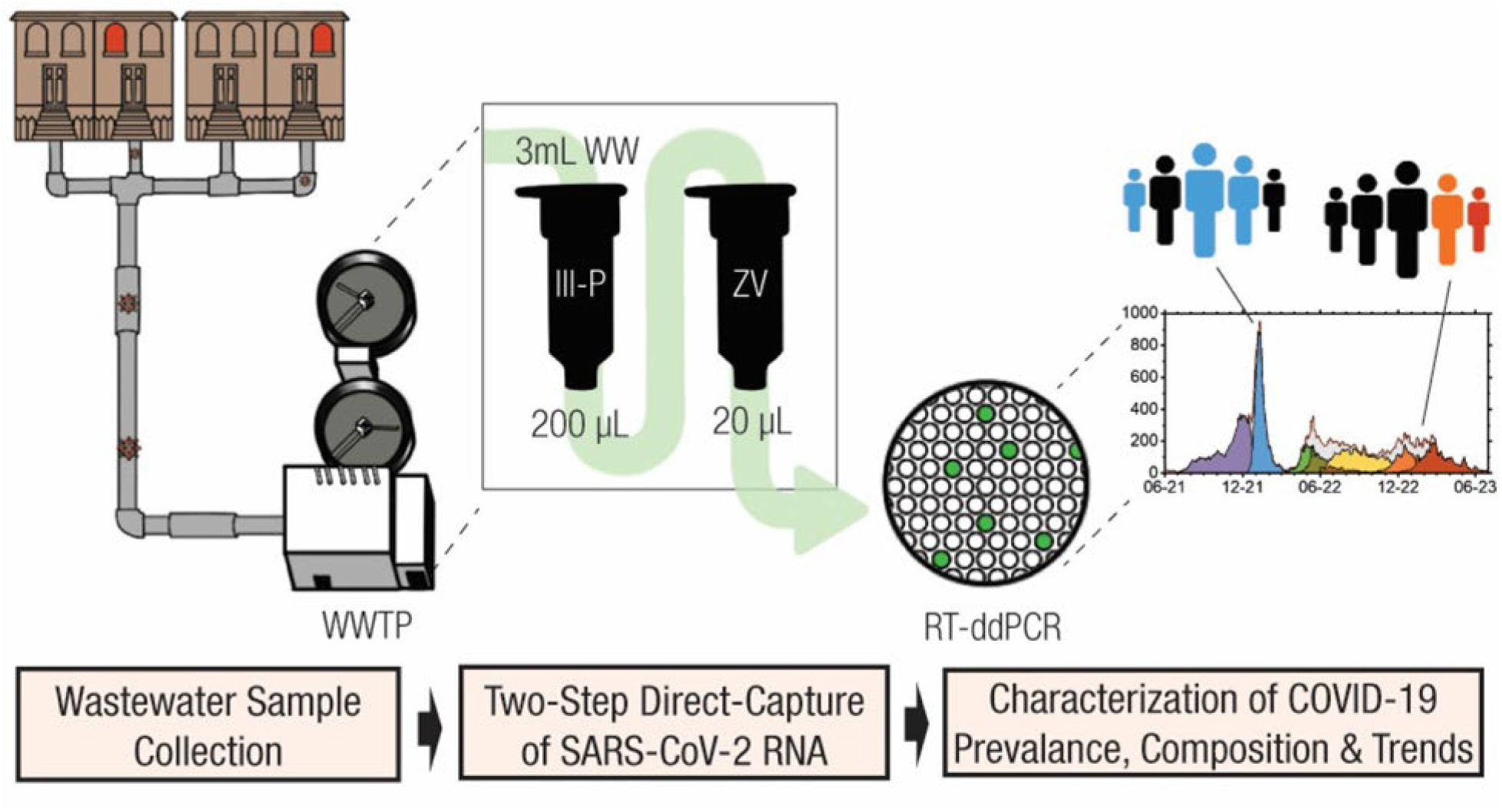

## 1. Introduction

Coronavirus disease 2019 (COVID-19) is caused by severe acute respiratory syndrome coronavirus 2 (SARS-CoV-2), a novel coronavirus that emerged in China in late 2019 and quickly spread around the world (Hu et al., 2021; da Silva et al., 2022). SARS-CoV-2 viral RNA is present in the stool of most people infected with COVID-19 and can be shed via this route for weeks after symptom onset (Natarajan et al., 2022; Arts et al., 2023). In sewered communities, this RNA enters the wastewater collection system and typically flows to a wastewater treatment plant (WWTP). Levels of the viral RNA measured there in the influent wastewater or primary sludge are correlated over time with the number of reported cases in the plant’s service area, often providing a leading indication of changing trends in reported clinical case numbers (Medema et al., 2020a; Ahmed et al.,2020a; Randazzo et al., 2020; La Rosa et al., 2020; Gonzalez et al.,2020; Wu et al., 2020; Peccia et al., 2020; Graham et al., 2021). In addition, viral RNA fragments in the wastewater and sludge carry mutations that are characteristic of the particular genomic variant strains of SARS-CoV-2 circulating in the served community (Crits-Christoph et al., 2021; Heijnen et al., 2021; Bar-Or et al., 2021; Lee et al., 2021; Yaniv et al., 2021; Swift et al., 2021; Karthikeyan et al., 2022; Lou et al., 2022; Wurtzer et al., 2022). The analysis of wastewater can provide a measure of both the amount and genomic diversity of SARS-CoV-2 present in the community, offering a comprehensive, cost-effective, and unbiased means of monitoring disease prevalence and genomic heterogeneity (Medema et al., 2020b; Tiwari et al., 2023; Parkins et al., 2023). The broad utility of wastewater-based surveillance for the monitoring of disease abundance and diversity in sewered communities has now been widely demonstrated, and efforts are underway to integrate multi-target pathogen surveillance programs into public health infrastructure in the U.S. and around the world (National Academies of Sciences, Engineering, and Medicine, 2023; Kirby et al., 2021; Keshaviah et al., 2023). With reduced clinical testing and the increased use of at-home tests, along with cutbacks in clinical sequencing initiatives to identify and quantify the spread of new variants, wastewater surveillance now occupies a primary role in providing timely information on trends in COVID-19 disease prevalence and variant diversity.

Here, we report data for daily SARS-CoV-2 RNA concentrations and loads entering a major metropolitan wastewater treatment plant over 32 months, from November 2020 through June 2023. In addition, we show results for analyses of specific targeted mutations in the viral RNA that distinguish the major variant sub-lineages circulating in the sewershed during this time. These data were obtained using a novel two-step direct-capture extraction method to isolate SARS-CoV-2 RNA from raw wastewater, in conjunction with reverse transcription-droplet digital polymerase chain reaction (RT-ddPCR) for specific RNA target detection. The primary objectives of the study were to: 1) Develop a sensitive and precise method for measuring SARS-CoV-2 viral RNA concentrations in raw wastewater; 2) Use this method to monitor SARS-CoV-2 RNA concentrations and total loads entering the Metropolitan Wastewater Treatment Plant (Metro Plant) in Saint Paul, MN, USA; and 3) Measure the prevalence of targeted mutations in the extracted SARS-CoV-2 RNA to characterize and monitor the presence, prevalence, and load of major genomic variants in wastewater entering the Metro Plant. The results demonstrate the utility of these methods to provide a reliable measure of the total load of SARS-CoV-2 RNA in wastewater treatment plant influent and chronicle the evolving variant composition in the plant’s service area.

## 2. Materials and Methods

### 2.1 Sample collection

Sampling was carried out at the Metro Plant in Saint Paul, Minnesota, USA, from November 1, 2020, through June 30, 2023. The plant treats 6.9 m^3^/sec of wastewater (2022 annual average) from 68 communities in the Twin Cities (Minneapolis-Saint Paul) metropolitan area, serving a total population of 1.95 million people, including everyone in both Minneapolis and Saint Paul. The Metro Plant is the largest of nine WWTP operated by the Metropolitan Council, the regional policy, planning, and service agency of the seven-county Twin Cities metropolitan area. In addition to the nine WWTP, the Metropolitan Council also owns and operates the separated sanitary interceptor sewer system that delivers wastewater to those plants.

Daily 24-hour flow-proportional composited raw wastewater samples were collected by refrigerated autosampler (4°C) from the primary influent channel at the Metro Plant (i.e., post-grit removal). The average, median, 10^th^ percentile, and 90^th^ percentile influent flow rates for the period of study were 7.0, 6.9, 6.5, and 7.6 m^3^/sec, respectively, and the range was 6.0 to 10.6 m^3^/sec. The autosampler was set up to collect one 65-70 mL subsample for every 4,807 m^3^ of wastewater entering the plant. The total number of sub-samples collected each day ranged from 109 to 190, and total daily sample volumes ranged from 7 L to 13 L. Samples were retrieved from the autosampler each morning, representing 24-hour samples collected from 6:00 AM to 6:00 AM.

All samples were stored at 4°C until further processing. A representative 250-mL subsample of the daily composite sample was homogenized at 12,000 rpm for three minutes. A 30-mL subsample of the homogenized sample was added to a 50-mL centrifuge tube containing 15 mL of Zymo DNA/RNA Shield 2X Concentrate (Zymo Research, Irvine CA, USA). The treated sample was repeatedly inverted to mix and then allowed to sit at room temperature for at least 75 minutes before proceeding. Most samples were processed in this way within one hour of retrieval of the daily sample from the autosampler; all samples were processed within 24 hours of retrieval.

### 2.2 RNA extraction

A 4.5-mL aliquot of the treated sample was combined with 12 mL 100% ethanol in another 50-mL centrifuge tube. This combination was mixed and then completely transferred to a Zymo-Spin III-P silica column with 50-mL reservoir (Zymo Research, CA) placed on a vacuum manifold. The sample was pulled through the column, and the flow-through was discarded. The column was washed first with 5 mL of wash buffer 1 (WB1: 1.5 M NaCl in 20% ethanol; Whitney et al., 2021) and then with 10 mL of wash buffer 2 (WB2: 0.10 M NaCl in 80% ethanol; Whitney et al., 2021), pulling the wash buffers by vacuum through the column in each case. The column reservoir was discarded, and the silica column was placed in a receiving tube and centrifuged at 13,000 rpm for 2 minutes to remove excess WB2. The column was placed in a 1.5-mL receiving tube, and a 200-μL aliquot of warm (50°C) ZymoPure Elution Buffer (Zymo Research, CA) was added to the column. The column was placed in a hot block at 50°C for 10 minutes, after which the RNA adsorbed on the silica column was eluted into the receiving tube by centrifugation at 13,000 rpm for 5 minutes.

The RNA in the 200-μL eluate from the first step was further cleaned and concentrated using the Zymo Quick-RNA Viral Kit (Zymo Research, CA). The entire 200-μL volume was mixed with 400 μL Viral RNA Buffer, and the 600-μL mixture was placed on a Zymo IC silica column (Zymo Research, CA). The column was centrifuged for 90 seconds to bind the RNA, and the flow-through was discarded. The column was then washed in three subsequent steps using 620 μL Zymo Viral Wash Buffer (VWB; Zymo Research, CA), 500 μL VWB, and 500 μL 100% ethanol, respectively. In each step, the wash liquid was pulled through the column by centrifugation at 13,000 rpm for 90 seconds, and the flow-through was discarded. The column was placed in a 1.5-mL receiving tube, and the RNA was eluted with 20 μL RNA-free pure water by centrifugation at 13,000 rpm for 1 minute.

For quality assurance, the concentration and quality (A260/A280) of the RNA in each extract was determined for each sample using the NanoDrop One UV-Vis spectrometer (Thermo Fisher Scientific, Waltham, MA, USA). RNA samples were stored at -80°C until analysis within one week by reverse transcription-droplet digital polymerase chain reaction (RT-ddPCR).

Daily wastewater influent samples were extracted in replicate, typically 6X or 8X. From 11/1/2020 to 6/20/2022, two of these daily extraction replicates were analyzed by RT-ddPCR (i.e., these are “method duplicates”). After 6/20/2022, the daily replicates were combined and then split into two separate subsamples for RT-ddPCR analysis (i.e., these are “ddPCR duplicates”). (“Method duplicates” are subsamples taken from the same daily sample and extracted separately; the extracts are not mixed but submitted for RT-ddPCR separately. The “ddPCR duplicates” are subsamples taken from the same daily sample and extracted separately; the extracts are then mixed and subsequently reseparated into two tubes and submitted for RT-ddPCR separately.)

### 2.3 Quantification of SARS-CoV-2 RNA by RT-ddPCR

Concentrations of SARS-CoV-2 RNA in wastewater sample extracts were determined by RT-ddPCR using assays targeting the SARS-CoV-2 nucleocapsid (N) gene at two loci, N1 and N2 (Lu et al., 2020). The relative abundances of specific SARS-CoV-2 RNA mutations in these samples were determined by RT-ddPCR assays targeting mutations of interest in the ORF1ab, ORF1b, ORF7b, S, and M regions of the viral genome. Reactions were prepared using the Bio-Rad One-Step RT-ddPCR Advanced Kit for Probes (Bio-Rad, Hercules, CA, USA) according to the kit protocol for both overall quantification and mutation assays.

Sample reaction mixtures were prepared as 20 μL, total volume, with 5 μL of template RNA and final concentrations of: 1x One-Step RT-ddPCR Advanced Kit for Probes, 20 U/μL Reverse Transcriptase, 15 mM dithiothreitol (DTT), 900 nM target primers, and 250 nM target probes. N1 and N2 assays were run as separate reactions, each consisting of primer with FAM-labelled probes. Mutation assays consisted of either FAM/HEX or FAM/VIC probes, where FAM was used to detect the mutant allele of the target gene (*Mutant*) and HEX or VIC was used to detect the wild-type allele (*WT*). Mutation assays were purchased from Bio-Rad and Applied Biosystems/Thermo Fisher Scientific (Waltham, MA, USA).

Negative and positive controls were included on each plate. No Template Controls (NTC) were made with RNase-/DNase-free water for all assays. Previously sequenced positive SARS-CoV-2 clinical samples and synthetic SARS-CoV-2 controls were used as positive controls.

Plates were sealed with Bio-Rad PX1 PCR Plate Sealer (Bio-Rad, CA) and sample reaction mixtures were partitioned into droplets using the Bio-Rad QX200 droplet generator (Bio-Rad, CA) with Bio-Rad Automated Droplet Generation Oil for Probes (Bio-Rad, CA), according to the manufacturer’s instructions.

RT-ddPCR cycling conditions for the N1, N2, and Thermo Fisher mutation assays: reverse transcription, 45°C (60 min); enzyme activation, 95°C (10 min); 40 cycles, each, of denaturation, 95°C (30 sec), and annealing, 60°C (1 min); enzyme deactivation, 98°C (10 min); and hold, 4°C (infinite), **(Table S1, Supplementary material)**. All temperature changes were performed with a ramp speed of 2°C/s per manufacturer recommendations.

Cycling conditions for the Bio-Rad mutation assays **(Table S1)**: reverse transcription, 45°C (60 min); enzyme activation, 95°C (10 min); 40 cycles, each, of denaturation, 95°C (30 sec), and annealing, 55°C (1 min); enzyme deactivation, 98°C (10 min); and hold, 4°C (infinite). The ramp speed was 2°C/s.

Following amplification, sample droplets were read using the Bio-Rad QX200 Droplet Reader (Bio-Rad, CA), and data was analyzed using Bio-Rad QuantaSoft™ Analysis Pro Software (Bio-Rad, CA). The QX200 Droplet Reader measures the intensity of fluorescence emissions (amplitude) of a reporter probe (FAM, HEX, or VIC) in each droplet.

Controls provided quantitative amplitude reference measurements for PCR-negative and PCR-positive droplets, from which an amplitude threshold level was determined. Sample droplets with an amplitude above this set threshold level were considered PCR-positive, and droplets below this threshold were considered PCR-negative. Total accepted droplet counts typically ranged from 16,000 to 19,000, and reactions with accepted counts less than 10,000 were excluded from analysis.

The fraction of PCR-positive droplets for each reaction was fit to a Poisson distribution model to determine the number of target gene fragments (copies/20 μL reaction). N1 and N2 results were reported as the concentration of target copies in the original RNA extract (copies/μL).

### 2.4 Mutation assays

Starting in June 2021, major variants present in Metro Plant influent were quantified by ddPCR using targeted mutation assays. For the targeted mutation assays, where *Mutant* and *WT* concentrations (copies/μL) were both determined in a single reaction, the relative abundance of targets containing the mutation (referred to here interchangeably as either “frequency” or “prevalence”) in a sample was calculated as:

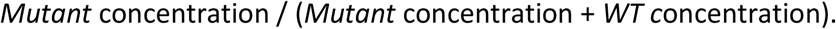

Details of the assays used are described in **Table S2**, and the calculations employed in estimating the variant frequencies and their periods of use are shown in **Table 1**. The calculation for some variant strains (e.g., Delta, BA.1, BA.4, BA.5, etc.) was based on a single mutation, unique to that strain. For others, the calculation utilized the results of two (e.g., BA.2, BA.2.12.1) or three (e.g., XBB) mutation assays. As indicated in the table, the calculations for some variants changed over the course of study as new mutation assays became available which we determined were more sensitive and/or specific to the variant we were tracking.

**Table 1.**
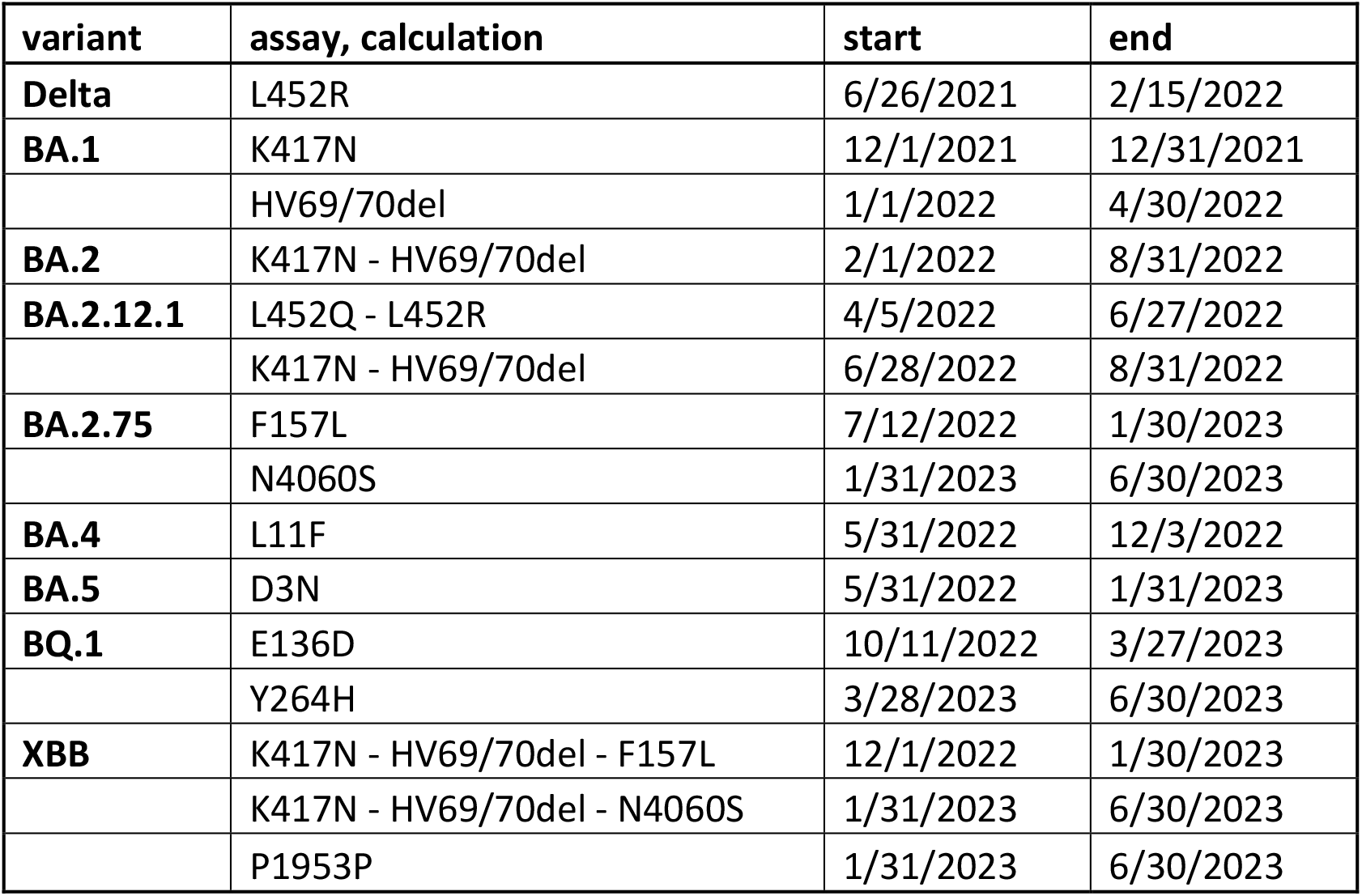
Timing of our use of various mutation assays, and calculations used to estimate variant prevalence in Metro Plant influent.

### 2.5 Dilution series

A low-level dilution series was prepared to illustrate the approximate lower limit of quantification of the analytical protocol. Aliquots of the influent sample from 4/18/21 were extracted in two separate batches. First, six aliquots were extracted, and the six extracts were combined and mixed well. The combined extract was diluted at 1:4, 1:16, 1:32, and 1:64 using RNase-free distilled water (Invirogen UltraPure DNase/RNase-Free Distilled Water, #10977023; Thermo Fisher Scientific, Waltham, MA), and triplicate samples at each level were prepared for RT-ddPCR analysis. In the second batch, six additional aliquots were extracted, and four of those extracts were combined and mixed well. This combined extract was diluted as before at 1:4, 1:16, 1:32, and 1:64, and quintuplicate samples at each level were prepared for RT-ddPCR analysis. The eight extract samples at each dilution level were analyzed for N1 and N2 by our standard RT-ddPCR protocol.

### 2.6 Extraction recovery

A sample of Metro Plant influent was spiked with Zeptometrix SARS-CoV-2 Extraction Run Control (ERC #325642; Zeptometrix, Buffalo, NY, USA) to quantify extraction recovery. This extraction control material consists of purified, noninfectious, intact viral particles in an aqueous solution of purified protein. A well-mixed aliquot (0.45 mL) of the ERC was added to an 18-mL subsample of influent in a 50-mL tube and mixed well by pipet and swirling. The mixture was immediately treated with 9 mL Zymo DNA/RNA Shield 2X (Zymo Research, CA) and mixed well by pipet. This procedure was then repeated two more times, resulting in 18 treated sub-samples for estimating the extraction recovery, each containing 3.0 mL of spiked wastewater. N1 and N2 concentrations in each sample were determined by RT-ddPCR and compared to the expected concentrations to estimate recovery. The extraction recovery was used only as a quality assurance indicator. N1 and N2 data were not corrected for extraction recovery.

### 2.7 Wastewater calculations

The population-normalized total daily load of SARS-CoV-2 RNA entering the Metro Plant was calculated by multiplying the daily influent flow rate by the measured concentration of viral RNA in the wastewater, then dividing that by 1.95E+06, the population in the Metro Plant service area.

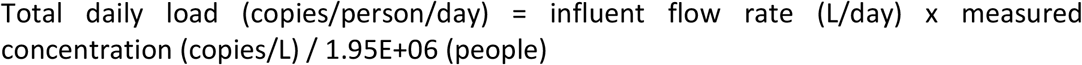

The concentration used here was the average of the N1 and N2 concentrations as determined by RT-ddPCR. The daily loads of individual variant sub-lineages were calculated by multiplying the total daily load by the frequency of the sub-lineage in that day’s sample.

### 2.8 Clinical case and sequencing data

Data for the number of new daily cases in Metro Plant service area zip codes were obtained from the Minnesota Department of Health (MDH). The total number of new daily cases within the Metro Plant service area was calculated by multiplying the GIS-derived estimate of fraction of the zip code area served by the Metro Plant by the total number of new daily cases in that area, summed over all zip code areas. Daily new case data is reported by specimen collection date.

Data for daily variant prevalence (frequency) in clinical specimens collected throughout Minnesota were provided by MDH. Select high-concentration clinical samples (PCR cycle threshold < 30) were contributed by COVID-19 testing sites and sequenced by the MDH Public Health Laboratory and other labs contracted by MDH or the Centers for Disease Control and Prevention (CDC) for this purpose. A total of 95,000 sequences collected between June 1, 2021, and June 30, 2023, were included in this analysis. Discrete data sets for the Delta, BA.1, BA.2 (excluding BA.2.12.1), BA.2.12.1, BA.5 (excluding BQ.1), BQ.1, and XBB variants were prepared for comparison with the corresponding wastewater data.

### 2.9 Statistical analysis

To determine the statistical correlation between wastewater and clinical data sets, the correlation statistics of the transformed, normalized datasets were quantified by standard Pearson correlation. Briefly, time-series datasets were ranked, and an inverse-normalization transformation was applied to each (Bishara and Hittner, 2012). Normalization results were checked using the Shapiro-Wilk test. We then calculated the Pearson correlation coefficient to quantify the correlation of daily wastewater SARS-CoV-2 load data with daily new clinical case data in the Metro Plant service area, and for the variant-specific correlation of daily variant frequency data in Metro Plant influent with that in sequenced clinical samples from across Minnesota.

## 3. Results and Discussion

### 3.1 Measuring total load in wastewater using a novel direct capture method

In October 2020, we developed a direct capture protocol for recovering SARS-CoV-2 RNA from raw wastewater. We coupled that with RT-ddPCR to characterize viral RNA concentrations in wastewater entering the Metro Plant in Saint Paul, MN. Daily monitoring began in November 2020 using only the Zymo *Quick*-RNA Viral Kit (#R1035, Zymo Research, CA) to extract RNA. This procedure was limited to a very small input volume (0.27 mL of wastewater), resulting in what we thought might possibly be an unacceptably high limit of quantification. In February 2021, aided by the work of Whitney et al. (2021), we added a preconcentration step to the extraction procedure to allow a larger wastewater volume (3 mL) to be extracted, followed then by the Zymo *Quick*-RNA Viral procedure. This change improved the sensitivity of our method, reducing the limit of quantification by as much as a factor of 3.0/0.27 = 11.1. The initial step involves a silica column-based extraction, just like the *Quick*-RNA Viral procedure, but uses a larger column with a 15-mL reservoir (#C1040-5, Zymo III-P Column Assembly; Zymo Research CA). The combined two-step procedure yields a method concentration factor of 150 (3 mL wastewater extracted into 20 uL final eluate). In practice, eight wastewater samples could be extracted within 90 minutes, requiring considerably less time than other reported methods for SARS-CoV-2 RNA concentration/extraction from wastewater (LaTurner et al., 2021).

The first known COVID-19 case in Minnesota was identified within the Metro Plant service area on March 5, 2020. The number of new daily cases reported in the service area varied through the summer of 2020, but remained below 600 until October of that year. Reported case numbers then increased rapidly before this wave peaked at 2657 new daily cases on November 9. Our monitoring of SARS-CoV-2 RNA in wastewater entering the Metro Plant in Saint Paul began on November 1, 2020, during the rapid increase in cases **(Figure 1)**. The population-normalized daily load of SARS-CoV-2 RNA in wastewater entering the plant tracked closely with the number of daily new cases in the Metro Plant service area throughout the full 32-month course of our study and was significantly correlated with daily new case counts (November 2020 through June 2023; n=970; r= 0.75, 95% CI [0.72, 0.77], p(α=0.05) <0.0001).

**Figure 1.**
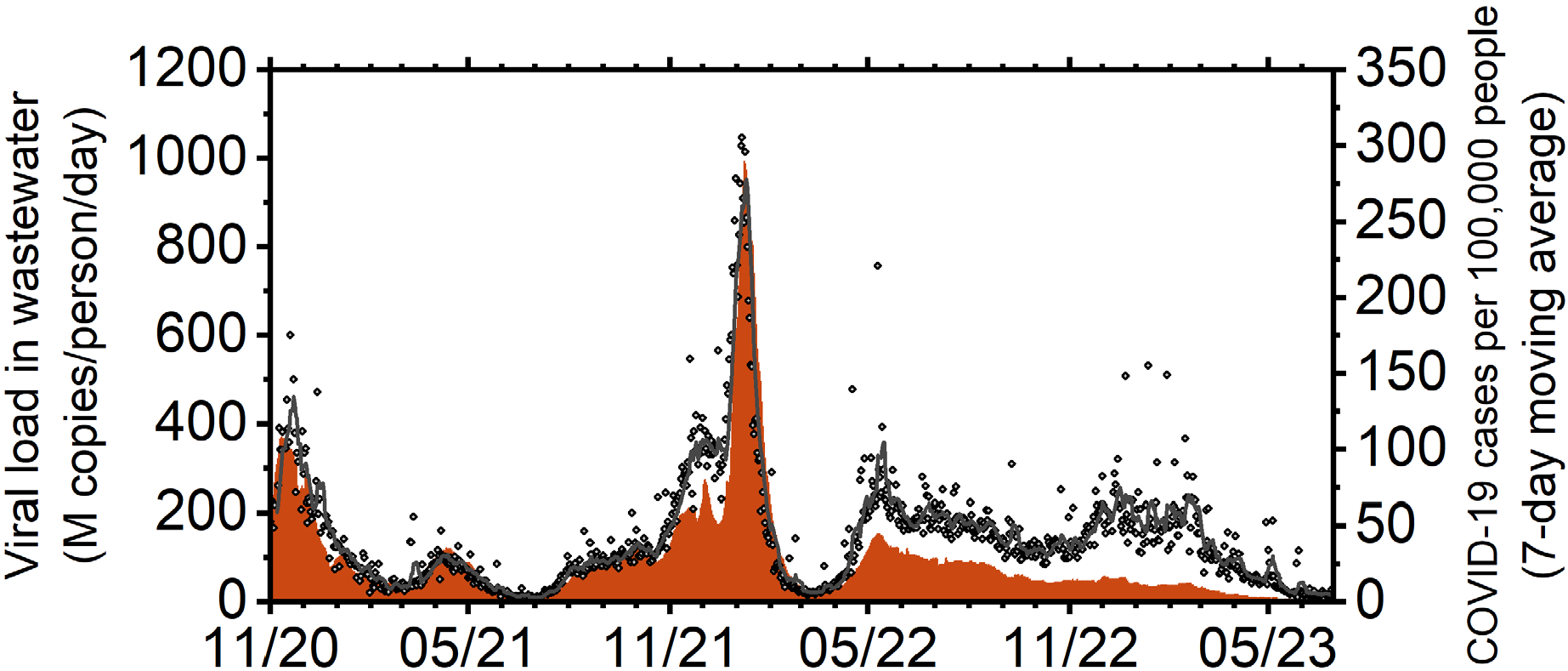
Each measured value of the total daily SARS-CoV-2 load in Metro Plant influent (diamonds) and the 7-day simple moving average (black line) of these data are plotted on the left, vs time. The 7-day simple moving average of the number of new daily clinical cases reported for the Metro Plant service area is plotted on the right in consolidated red bars, vs time.

N1 and N2 concentrations were quantifiable in all daily samples of Metro Plant influent collected over the course of this study. We used the average of the N1 and N2 concentrations to calculate the total load of SARS-CoV-2 RNA entering the Metro Plant. The average (N1,N2) concentration in Metro influent ranged from a low of 22,608 copies/L on 6/21/21 to a high of 3,548,708 copies/L on 1/6/22.

Multiple (typically 6-8) aliquots of treated sample were extracted for each daily sample. Two of those extracts were submitted for RT-ddPCR for each day between 11/1/20 and 6/20/22 (these are “method duplicates”). The average relative standard deviation of these 496 method duplicate pairs was 12% for N1 and 11% for N2. On and after 6/21/22, the replicate extracts from each daily sample were mixed together and then re-separated into separate tubes, and two of those tubes were submitted for RT-ddPCR (these are “ddPCR duplicates”). The average relative standard deviation of these 429 ddPCR duplicate pairs was 10% for N1 and 8% for N2. These results indicate that RT-ddPCR analysis is responsible for most of the overall method variability, with extraction introducing considerably less uncertainty (approximately 2-3%).

Our dilution series analysis of a single influent sample showed that N2 was detected in all eight of the most-diluted sub-samples (average = 6,300 copies/L, relative standard deviation = 35%, n=8), while N1 was detected in seven of the eight most-diluted samples (average = 4,300 copies/L, relative standard deviation = 37%, n=7). Data are shown in **Table S3** and **Figure S1**. Overall, the dilution study results suggest that our method could quantify SARS-CoV-2 RNA in wastewater at concentrations as low as approximately 5000 copies/L, well below the lowest concentration we observed (average (N1,N2) = 22,608 copies/L). Our ddPCR assay for N2 was more sensitive than that for N1, and the measured N2 concentrations were routinely higher than the N1 concentrations measured in the same sample.

A SARS-CoV-2 viral RNA extraction control was spiked into Metro Plant influent wastewater, and 18 samples were extracted separately to determine the recoveries of N1 and N2 gene fragments **(Table S4)**. The mean recovery of N1 was 84% (relative standard deviation (RSD) = 18%; range = 66% - 122%; n=18), and that of N2 was 86% (RSD = 15%; range = 62% - 115%; n=18). These results illustrate the extraction recovery performance of our method only for this particular wastewater sample. We did not correct daily N1 and N2 results for extraction recovery.

As has been reported in other locations, we observed successive waves of increased infections evidenced in both the number of reported cases in the Metro Plant service area and in viral RNA levels in Metro Plant influent **(Figure 1)**. After the large number of infections observed in late 2020, a smaller wave was observed in the spring of 2021, associated with the local circulation of the Alpha variant. A lull in local viral activity through early summer 2021 resulted in the lowest levels of SARS-CoV-2 RNA in Metro Plant influent we observed, with a minimum load observed on June 26. An incipient new wave driven by the introduction of the Delta variant followed quickly and continued through December 2021. Much higher viral RNA levels were observed in Metro influent in January 2022 as the Omicron BA.1 variant swept through the local region, with a maximum load observed on January 6. The viral load entering Metro remained relatively high through subsequent waves of Omicron sub-lineages, from May 2022 through February 2023, but the number of reported cases decreased almost continuously over that period.

The excellent correlation of the measured wastewater SARS-CoV-2 load data with the reported daily case data for the Metro Plant service area suggests that our method provides an accurate and reliable measure of local viral circulation. Since the start of the COVID-19 pandemic, many studies have demonstrated the value of wastewater surveillance for tracking community levels and trends of the SARS-CoV-2 virus (Tiwari et al., 2023; Parkins et al., 2023). A variety of approaches have been employed to isolate and quantify SARS-CoV-2 RNA from wastewater, and several effective methods have been reported (Pecson et al., 2021). Because of the relatively low concentrations of viral RNA in wastewater, some type of concentration step has often been employed to capture sufficient RNA for subsequent analysis. Concentration methods reported include electronegative membrane filtration, precipitation, ultrafiltration, and ultracentrifugation (Ahmed et al., 2020b; LaTurner et al., 2021; Zheng et al., 2022; Farkas et al., 2022). Recovery of viral RNA using these techniques was often found to be low and sometimes variable, however, introducing additional uncertainty into the observed viral concentration data (Pecson et al., 2021; LaTurner et al., 2021; Zheng et al., 2022).

Several previous studies have, like ours, taken a completely different approach, choosing instead to lyse the viral particles in the wastewater sample, stabilize the freed RNA, and concentrate the RNA during the extraction procedure (Whitney et al., 2021; Mondal et al., 2021; Song et al., 2021; Ferreira et al., 2022; Girón-Guzmán et al., 2023; Gupta et al., 2023; Haeusser et al., 2023). These “direct capture” methods can be applied to the whole wastewater sample (i.e., liquid plus solids) and benefit from their avoidance of time- and resource-consuming concentration steps, the poor and potentially variable recovery outcomes associated with those steps, and no loss of sensitivity due to preferential treatment of one sample fraction over another. Compared to methods employing a concentration step, direct capture methods may process relatively small initial volumes of wastewater and, thus, produce less RNA for ultimate detection, but their sensitivity can be sufficient to track SARS-CoV-2 RNA in wastewater at levels well below even the lowest levels seen in many communities.

With a concentration factor of 150, our “direct capture” method was sufficiently sensitive to quantify even the lowest RNA concentrations in wastewater entering the Metro Plant over 32 months of monitoring. The extraction method produced very high-quality RNA (A260/A280 > 2.0), and the approximate minimum level of quantification was found to be around 5000 copies/L, translating to a minimum measurable daily load of <2 million copies/person/day. Furthermore, the extraction method was fast and straightforward compared to many other reported wastewater extraction methods and introduced very little additional variability (approximately 2-3%) into the resulting data. With both time saving and data quality advantages, direct capture methods offer attractive benefits for pursuing wastewater surveillance of SARS-CoV-2 and other viral targets.

Our analytical workflow coupled a “direct capture” extraction procedure with droplet digital PCR. Many reported studies of SARS-CoV-2 RNA in wastewater have utilized quantitative polymerase chain reaction (qPCR) to quantify select gene fragments in the sample extracts. Others have preferred digital PCR (dPCR) platforms (Ciesielski et al., 2021; Feng et al., 2021; Flood et al., 2021), which offer improved accuracy, precision, and sensitivity compared to qPCR, as well as absolute quantitation, thereby eliminating the external standardization required for qPCR (Ahmed et al., 2022; Tiwari et al., 2022, Ding et al. 2024). And while qPCR reactions can suffer inhibition from the effects of sample contamination, dPCR is less susceptible to inhibition of the PCR reaction because it uses highly diluted reactions and endpoint fluorescence measurement. The high sensitivity and other advantages of dPCR offer the opportunity to explore the presence and prevalence of SARS-CoV-2 variants at low concentrations in wastewater, providing a quantitative picture of population-level variant diversity and prevalence in the served community.

### 3.2 Variant prevalence in wastewater

We exploited the exceptional target sensitivity of ddPCR to quantify low-abundance SARS-CoV-2 variant alleles in the complex RNA samples extracted from Metro Plant wastewater. The sequencing of wastewater-derived SARS-CoV-2 RNA has now been widely reported, demonstrating the capability of this approach for characterizing genomic diversity (Crits-Christoph et al., 2021; Bar-Or et al., 2021; Swift et al., 2021; Karthikeyan et al., 2022) and often showing remarkable agreement between variant prevalence signals in the wastewater and clinical data sets (Radu et al., 2022; Agrawal et al., 2022; Jahn et al., 2022; Izquierdo-Lara et al., 2023; Gupta et al., 2023). The sequencing of complicated samples like those derived from wastewater can be costly and time-consuming, however, requiring advanced instrumentation, complex bioinformatic analysis, and the necessary expertise to carry out the work. Sequencing workflows typically take more than a week to complete. Alternatively, assaying targeted mutations in the SARS-CoV-2 genome using dPCR or qPCR offers a quicker and cheaper approach to characterize viral variant diversity in wastewater (Heijnen et al., 2021; Lee et al., 2021; Yaniv et al., 2021; Lou et al., 2022; Wurtzer et al., 2022; Tiwari et al., 2023). In this case, a single mutation uniquely specific to a particular variant can be targeted for analysis, or multiple mutations can be quantified that, together, uniquely identify a variant. Appropriate assays can be obtained from commercial vendors or designed in-house, and results can accurately describe the relative abundance of the targeted alleles even within a complicated mixture of variants. Digital PCR platforms are capable of accurately and precisely quantifying low-frequency genomic mutations with high sensitivity and, thus, perfectly suited for characterizing SARS-CoV-2 variants in wastewater samples containing many distinct variant sub-lineages.

We used targeted mutation assays with RT-ddPCR to quantify individual variant alleles in Metro Plant wastewater (**Tables 1 and S2**). We began daily monitoring of the L452R mutation for the Delta variant in July 2021 and incorporated new targeted mutation assays into our analytical scheme over the remainder of the study to track the changing array of major variants circulating in the Metro Plant service area (see Table 1). The prevalence of each individual variant in wastewater generally tracked that observed in statewide clinical testing at the time, over the remaining course of the study **(Figure 2)**. Variant prevalence in Metro Plant wastewater was significantly correlated with that in sequenced clinical samples across all variants monitored from June 2021 through June 2023 (p (α=0.05) <0.00001; **Table S5**).

**Figure 2.**
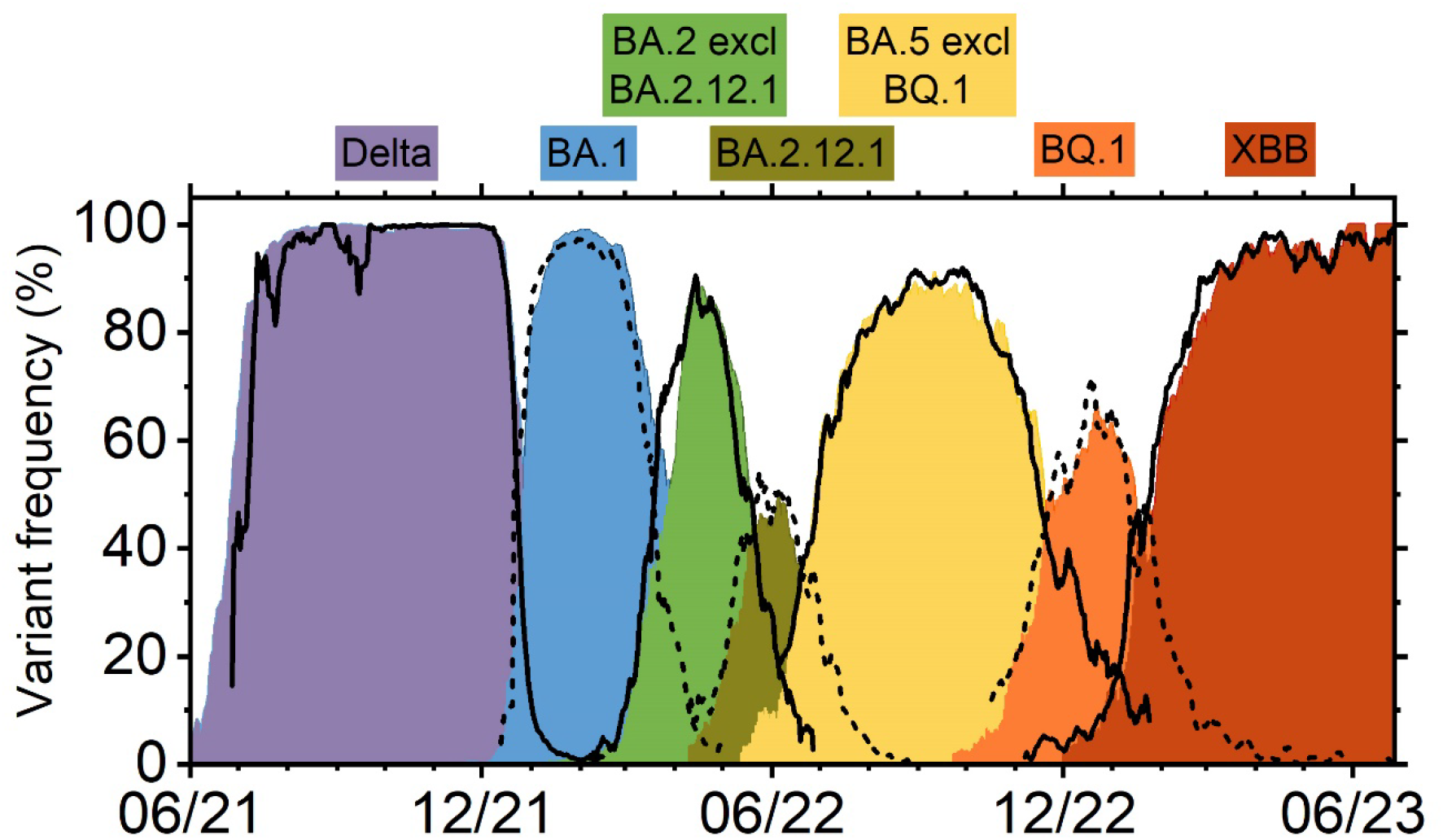
The 7-day simple moving average of the frequency of individual variants in clinical sequences collected in Minnesota is shown as consolidated colored bars, vs time. The 7-day simple moving average of the frequency of individual variants in Metro Plant influent is shown in solid or dashed black lines, vs time. Variants are labeled across the top.

Despite the overall close correlation of Metro Plant wastewater variant data with clinical test data over time, there was a noticeable discrepancy between the BA.2 frequency observed in wastewater and that reported by clinical testing in March and April 2022, when BA.2 was replacing BA.1 locally **(Figure 2)**. This discrepancy was traced to atypical clinical test data reported by a single contract laboratory (“Lab A”) for samples collected between March 21 and April 2. Of the 565 clinical samples collected from across Minnesota during this 13-day period and subsequently sequenced, 147 originated from Lab A, and 418 originated from the 40 other contributing laboratories. A total of 288 BA.2 sequences were found during this period among the 418 samples originating with labs other than Lab A, for an average BA.2 prevalence of 0.69. Of the 147 samples from Lab A, however, only two were reported as BA.2, resulting in an average BA.2 prevalence of 0.01. For comparison, the average BA.2 prevalence measured in Metro Plant wastewater over this period was 0.73.

The samples analyzed by both Lab A and the 40 other labs originated from locations similarly distributed across Minnesota, suggesting that similar populations were sampled. As a result, we expected the BA.2 prevalence in the two data sets to be similar, but it was not. We were unable to determine a reason for the apparent shortfall of BA.2 sequences in Lab A samples from this period, and thus included the Lab A data in our analysis here. Omitting the atypical Lab A data leads to much closer agreement between the wastewater and clinical BA.2 prevalence data in March/April 2022 **(Figure S2)**.

In April 2022, we started using the L452Q mutation assay to estimate the BA.2.12.1 frequency in wastewater. Phylogenetic analysis of clinical sequences had indicated that the L452Q mutation was essentially exclusive to the BA.2.12.1 variant when it appeared in Minnesota in April 2022, and we expected the L452Q mutation frequency would reflect the BA.2.12.1 frequency. By May, however, we started to see that the L452Q frequency in wastewater was reading higher than that in clinical sequencing results for BA.2.12.1. We hypothesized that the L452Q assay might also be reporting positive for another emerging mutation at that same locus, L452R, which started increasing in Minnesota by the end of April 2022 due to initial increases of BA.4 and BA.5, both of which carry that mutation. We were simultaneously measuring L452R, also, and found we could accurately track the clinical BA.2.12.1 frequency by subtracting the L452R allele frequency from that determined by the L452Q assay. The dashed line showing the wastewater frequency for BA.2.12.1 in **Figure 2** is for “L452Q minus L452R,” and it follows the clinical results quite well. These results suggest that the mutant allele primers for the L452Q assay were not specific to detection of the Q substitution at site 452, but positively amplified the L452R mutation as well. The L452R assay, on the other hand, was specific to the L452R mutation and unaffected by the L452Q mutation in the S-gene.

Overall, these results show that using targeted mutation assays in combination with RT-ddPCR is an effective strategy for tracking SARS-CoV-2 viral variants in wastewater. The samples extracted for monitoring N1 and N2 gene targets were well suited, also, for providing timely and accurate variant data using the targeted mutation/ddPCR approach. We used commercially available mutation assays, and our project was thus constrained by the availability of the appropriate assays. This limited our work to tracking only previously identified variants, but that was not a severe constraint since assays for the variants of interest to us were typically available by the time we needed them. Our approach generated accurate variant prevalence data for our wastewater samples in a timely and cost-effective manner. Also, remarkably, our results demonstrate that monitoring the variant composition and diversity in wastewater at the state’s largest wastewater treatment plant in Saint Paul, serving only 34% of the total Minnesota population, served to provide an accurate measure of variant prevalence over time for the entire state population.

### 3.3 Calculated daily and cumulative loads of variants in wastewater

We calculated the daily load of each variant entering the Metro Plant by multiplying the total daily load by the frequency observed for that variant on that day **(Figure 3)**. We estimated the total cumulative amount of all SARS-CoV-2 viral RNA entering the Metro Plant from June 1, 2021, through June 30, 2023, by summing the measured daily total loads observed over this period. Similarly, the cumulative amounts of each variant were calculated by summing the measured daily loads for that variant over this period.

**Figure 3.**
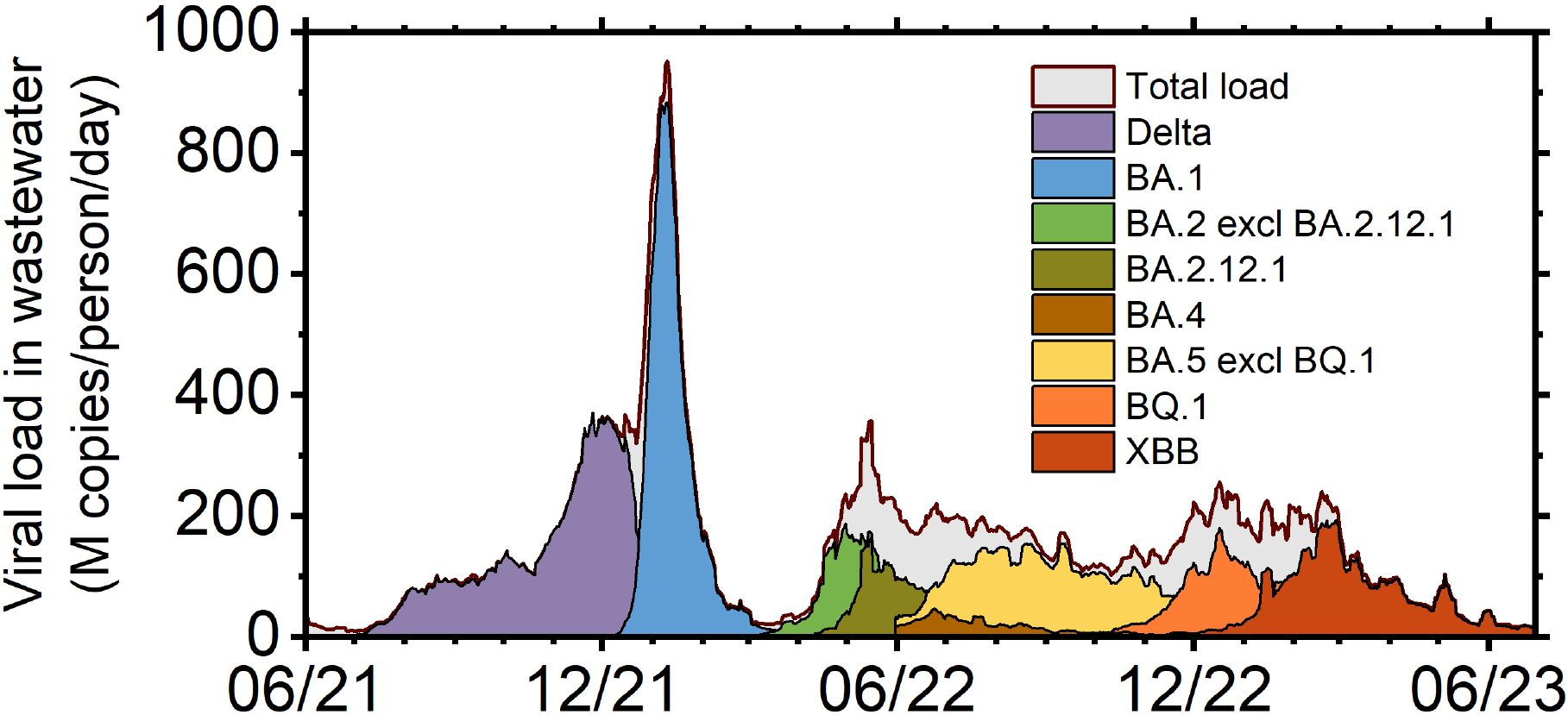
The 7-day simple moving average of the load of individual variants in Metro Plant influent is shown in consolidated colored bars with black outline, vs time, from June 1, 2021, to June 30, 2023. The 7-day simple moving average of the total daily SARS-CoV-2 load in Metro Plant influent is shown in consolidated gray bars with red/black outline, vs. time.

The total amounts of the various variant sublineages entering the plant and their relative proportions are shown in **Table S6**. A total of 2.30E+17 copies of the viral RNA entered the plant during this time (=117,800 million copies/person), and BA.5.X (27% of the total), BA.2.X (26%), and Delta (22%) were the largest contributing variant families. The BA.5.X lineage included BQ.1.X (8% of the total), and the BA.2.X lineage included BA.2.12.1 (6% of the total), BA.2.75 (1%), and XBB (12%). BA.4.X accounted for only 2% of the total amount of RNA we measured. BA.1.X registered the highest daily loads observed during our study, but overall, BA.1 and its sub-lineages accounted, cumulatively, for only 19% of the total amount of SARS-CoV-2 RNA entering the plant from June 2021 through June 2023.

Summing all the variant cumulative loads showed that our variant surveillance detected 96% of the total RNA entering the plant; i.e., only 4% of the viral RNA entering the plant during this time was from a variant we were not monitoring **(Figure S3)**. This small discrepancy may reflect the presence of unmeasured minor variants in the influent, but also reflects the uncertainty in our measurements and differences in the performance of the assays used for quantifying the total load (N1 and N2) and the variant loads.

## 4. Conclusions

We developed a fast and straightforward two-step, column-based direct capture extraction method and applied it, along with RT-ddPCR, to reliably quantify the total amount and genomic diversity of SARS-CoV-2 RNA in wastewater entering a large wastewater treatment plant in Saint Paul, MN, USA, over a 32-month period. The results show that the total amount of RNA in the wastewater was a dependable indicator of the levels of clinical cases reported in the treatment plant’s service area while clinical testing was widespread, and trends in the clinical case numbers were accurately reflected in the wastewater signal, providing timely information on the local disease situation and trends to the community. After widespread clinical testing declined, the wastewater data remained a reliable indicator of local disease prevalence and trends. The prevalence of major variants circulating in the community, as indicated by statewide clinical testing, was reliably quantified in the wastewater using targeted mutation assays and RT-ddPCR, providing public health authorities with rapid and low-cost confirmation of the clinical testing data and an alternative indicator of local variant prevalence as clinical sequencing decreased. Overall, the methods reported here offer a simple, reliable, and effective approach to tracking COVID-19 disease levels and variant prevalence in a community through wastewater surveillance.

## Data Availability

SARS-CoV-2 RNA load and variant prevalence data are available at https://metrotransitmn.shinyapps.io/metc-wastewater-covid-monitor/

## Supplementary material

Supplementary material associated with this paper can be found in the online version.

Table S1: RT-ddPCR conditions; Table S2: Targeted mutation assays used in this study; Table S3. Dilution series data. Table S4: Recovery of SARS-CoV-2 RNA from spiked wastewater; Table S5: Pearson correlation coefficients, r, of RIN-transformed data sets of 1) daily total wastewater load with daily new case counts; and 2) daily variant prevalence in wastewater with daily variant prevalence in MDH clinical sequences; Table S6: Total amounts of total and variant RNA entering the Metro Plant from June 1, 2021, through June 30, 2023; Figure S1: Dilution series results showing the average N1 and N2 concentrations (n=8) at each dilution level; Figure S2. Variant frequency data with clinical data submitted by “Lab A” from March 21 to April 2, 2022, omitted. Figure S3: Stacked area plot of variant loads over time.

## Author Contributions

**CRediT: Steven J. Balogh:** Conceptualization, Data curation, Formal analysis, Funding acquisition, Investigation, Methodology, Project administration, Resources, Supervision, Validation, Visualization, Writing – original draft, Writing – review & editing. **George B. Sprouse:** Conceptualization, Formal analysis, Funding acquisition, Project administration, Supervision, Writing – review & editing. **Kenneth B. Beckman:** Conceptualization, Methodology, Writing – review & editing. **Ray H. B. Watson:** Conceptualization, Data curation, Formal analysis, Investigation, Methodology, Resources, Supervision, Validation, Writing – review & editing. **Darrell M. Johnson:** Data curation, Investigation, Methodology, Resources, Supervision, Validation. **Lee Pinkerton:** Conceptualization, Investigation, Writing – review & editing. **Yabing H. Nollet:** Conceptualization, Investigation, Writing – review & editing. **Adam W. Sealock:** Conceptualization, Investigation, Writing – review & editing. **Walter S.C. Atkins:** Conceptualization, Visualization, Writing – review & editing. **Laura M. Selenke:** Data curation, Investigation. **Joseph A. Kinney:** Data curation, Investigation, Writing – review & editing. **Patrick J.R. Grady:** Data curation, Formal analysis, Investigation, Methodology, Resources, Supervision, Validation. **Brandon Vanderbush:** Data curation, Investigation. **Jerry J. Daniel:** Resources, Supervision.

## Acknowledgements

This work was funded internally by Metropolitan Council Environmental Services (MCES) in 2020 and 2021. Work in 2022 and 2023 was financed by American Rescue Plan funds provided by the State of Minnesota.

Special thanks to Bonnie Kollodge, Crystal Mulry, Laura Fletcher, Liz Roten, and Ashley Asmus at MCES for their contributions to the reporting of these data via our public dashboard.

We’re grateful, also, to: Monica Rose and MCES Laboratory staff for the prompt daily preparation of 250-mL wastewater subsamples for our use; Steve Hack and Meghan Wilson at MCES for GIS estimates of zip code area fractions in the Metro Plant service area; Kristin Sweet and Genny Grilli at the Minnesota Department of Health (MDH) for providing zip-coded daily case data; Scott Seys and other MDH staff for providing variant-specific clinical case data; and Daniel Huff, Ruth Lynfield, Beth Gyllstrom, Myra Kunas, and Sara Vetter at the Minnesota Department of Health, Mark Osborn at the University of Minnesota School of Medicine, and Michael Osterholm at the University of Minnesota School of Public Health for helpful discussions and encouragement.

