## Supplementary material for "Long-term monitoring of SARS-CoV-2 load and variant composition at a large metropolitan wastewater treatment plant using a simple two-step direct capture RNA extraction, droplet digital PCR, and targeted mutation assays"

### Abstract

Wastewater surveillance offers an objective, comprehensive, and cost-effective means of monitoring the prevalence and genomic heterogeneity of pathogens circulating in a community. Here, a novel two-step extraction procedure for the direct capture of SARS-CoV-2 RNA from raw wastewater is presented. Combined with reverse transcription-droplet digital polymerase chain reaction (RT-ddPCR) detection, the method provides a fast and sensitive method for measuring viral RNA concentrations in wastewater. The method was used to measure the concentration of SARS-CoV-2 RNA in daily samples of wastewater entering a major metropolitan wastewater treatment plant over the course of 32 months, from November 2020 through June 2023. In addition, targeted mutation assays were used with RT-ddPCR to characterize the evolving presence and prevalence of specific SARS-CoV-2 variant sub-lineages in the wastewater stream over time. The results demonstrate the utility of these methods to accurately measure the total load of SARS-CoV-2 RNA, and chronicle its evolving variant composition, in wastewater treatment plant influent, providing near-real-time characterization of COVID-19 disease prevalence and trends in the served community.

**Contents:**

Table S1: RT-ddPCR conditions.

Table S2: Targeted mutation assays used in this study.

Table S3. Dilution series data.

Table S4: Recovery of SARS-CoV-2 RNA from spiked wastewater.

Table S5: Pearson correlation coefficients, r, of RIN-transformed data sets of 1) daily total wastewater load with daily new case counts; and 2) daily variant prevalence in wastewater with daily variant prevalence in MDH clinical sequences.

Table S6: Total amounts of total and variant RNA entering the Metro Plant from June 1, 2021, through June 30, 2023.

Figure S1: Dilution series results showing the average N1 and N2 concentrations (n=8) at each dilution level.

Figure S2. Variant frequency data with clinical data submitted by "Lab A" from March 21 to April 2, 2022, omitted.

Figure S3: Stacked area plot of variant loads over time.

### Table S1. RT-ddPCR conditions.

| Cycling step | N1, N2, and Thermo Fisher Mutation Assays | Bio-Rad Mutation Assays | Time | Number of cycles |
|---|---|---|---|---|
| | Temperature °C | | | |
| **Reverse transcription** | 45 | 45 | 60 min | 1 |
| **Enzyme activation** | 95 | 95 | 10 min | 1 |
| **Denaturation** | 95 | 95 | 30 sec | 40 |
| **Annealing/extension** | 60 | 55 | 1 min | 40 |
| **Enzyme deactivation** | 98 | 98 | 10 min | 1 |
| **Hold** | 4 | 4 | infinite | 1 |
| **Ramp rate for all temperature changes = 2°C/sec** | | | | |

### Table S2. Targeted mutation assays used in this study.

| mutation | vendor | assay number |
|---|---|---|
| **S.L452R** | Bio-Rad | dMDS983315944 |
| **S.HV69/70del** | Bio-Rad | dMDS944624402 |
| **S.K417N** | Bio-Rad | dMDS817055273 |
| **S.L452Q** | Thermo Fisher | CVKA3AV |
| **ORF7b.L11F** | Thermo Fisher | CVCE3VH |
| **M.D3N** | Thermo Fisher | CVAAAAK |
| **N.E136D** | Thermo Fisher | CV9HHWW |
| **ORF1b:Y264H** | Thermo Fisher | CV47VR3 |
| **S.F157L** | Thermo Fisher | CVGZE4Y |
| **ORF1ab.N4060S** | Thermo Fisher | CVCE3VJ |
| **ORF1b.P1953P** | Thermo Fisher | CV32Z67 |

**Table S3. Dilution series data. Samples A1-A8 are at 1:4 dilution; B1-B8 are at 1:16 dilution; C1-C8 are at 1:32 dilution; and D1-D8 are at 1:64 dilution.**

| | N1 | N2 |
|---|---|---|
| sample | cp/L WW | cp/L WW |
| A1 | 54556 | 63029 |
| A2 | 67175 | 50597 |
| A3 | 61164 | 50295 |
| A4 | 67077 | 73053 |
| A5 | 70719 | 86930 |
| A6 | 58961 | 78971 |
| A7 | 45850 | 64115 |
| A8 | 62411 | 73810 |
| B1 | 13402 | 20525 |
| B2 | 16001 | 12399 |
| B3 | 13099 | 11043 |
| B4 | 14185 | 13889 |
| B5 | 11624 | 32302 |
| B6 | 11836 | 17947 |
| B7 | 18008 | 17150 |
| B8 | 13933 | 30231 |
| C1 | 9559 | 10911 |
| C2 | 5581 | 11548 |
| C3 | 7321 | 7802 |
| C4 | 5438 | 14031 |
| C5 | 5608 | 17380 |
| C6 | 6970 | 14146 |
| C7 | 7213 | 10917 |
| C8 | 6259 | 6164 |
| D1 | 3476 | 8265 |
| D2 | 1843 | 3312 |
| D3 | 5483 | 7515 |
| D4 | 5676 | 7025 |
| D5 | 0 | 8924 |
| D6 | 3420 | 4872 |
| D7 | 4998 | 6988 |
| D8 | 4964 | 3146 |

**Table S4. Recovery of SARS-CoV-2 RNA from spiked wastewater (%).**

| | N1 | N2 |
|---|---|---|
| **1** | 87% | 62% |
| **2** | 79% | 80% |
| **3** | 111% | 106% |
| **4** | 75% | 83% |
| **5** | 79% | 96% |
| **6** | 77% | 85% |
| **7** | 82% | 101% |
| **8** | 74% | 87% |
| **9** | 122% | 80% |
| **10** | 86% | 95% |
| **11** | 74% | 83% |
| **12** | 76% | 88% |
| **13** | 74% | 80% |
| **14** | 105% | 115% |
| **15** | 98% | 87% |
| **16** | 73% | 84% |
| **17** | 70% | 75% |
| **18** | 66% | 69% |
| **avg** | **84%** | **86%** |
| **stdev** | **15%** | **13%** |
| **%CV** | **18** | **15** |

**Table S5. Pearson correlation coefficients, r, of RIN-transformed data sets of 1) daily total wastewater load with daily new case counts; and 2) daily variant prevalence in wastewater with daily variant prevalence in MDH clinical sequences; n = number of daily data pairs; p = p-value at α=0.05.**

| statistical comparison | r | n | p @ α=0.05 |
|---|---|---|---|
| 1) total load vs. daily new cases | 0.75 | 970 | <0.00001 |
| 2) variant prevalence, wastewater vs. clinical | | | |
| Delta | 0.75 | 211 | <0.00001 |
| BA.1 | 0.89 | 144 | <0.00001 |
| BA.2 excluding BA.2.12.1 | 0.84 | 117 | <0.00001 |
| BA.2.12.1 | 0.88 | 116 | <0.00001 |
| BA.5 excluding BQ.1 | 0.87 | 235 | <0.00001 |
| BQ.1 | 0.85 | 161 | <0.00001 |
| XBB | 0.78 | 192 | <0.00001 |

**Table S6. Total amounts of total and variant RNA entering the Metro Plant during our variant monitoring effort (i.e., from June 1, 2021, through June 30, 2023).**

| | sum (M copies/person) | % of total |
|---|---|---|
| **Total RNA** | **117795** | |
| Delta | 26105 | 22% |
| BA.1 | 22229 | 19% |
| BA.2.12.1 | 6759 | 6% |
| BA.2 excl BA.2.12.1 | 8000 | 7% |
| BA.4 | 2902 | 2% |
| BQ.1 | 9782 | 8% |
| BA.5 excl BQ.1 | 21422 | 18% |
| BA.2.75 | 1227 | 1% |
| XBB | 14452 | 12% |
| **sum of measured variants** | **112878** | **96%** |
| **other (unmeasured)** | | **4%** |

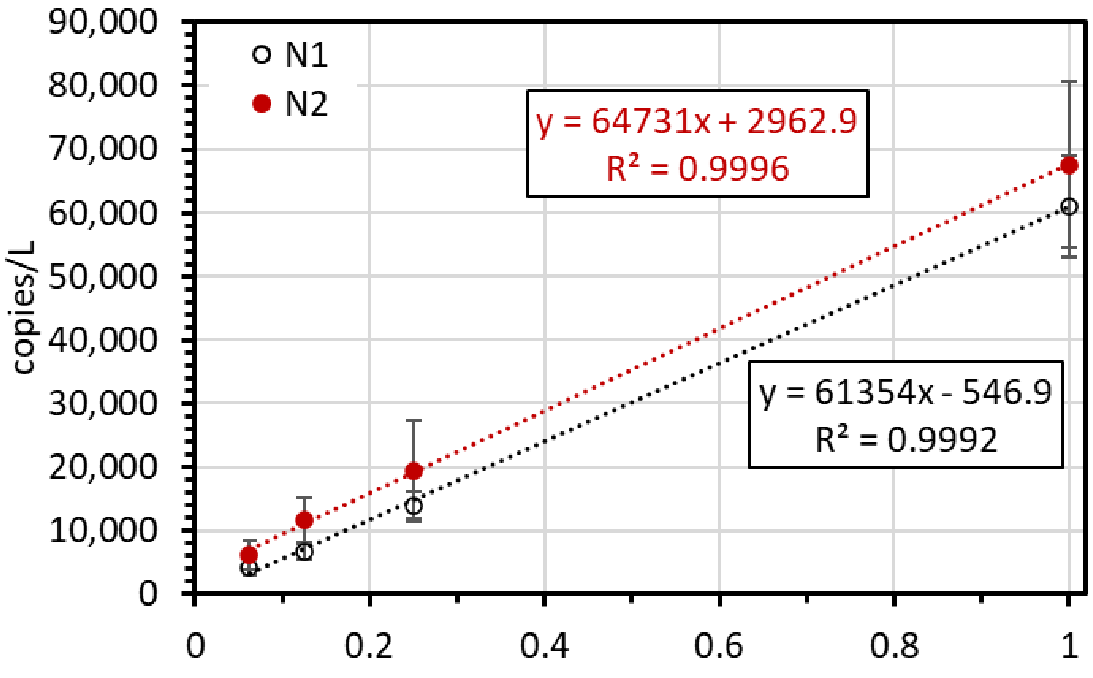


**Figure S1. Dilution series results showing the average N1 and N2 concentrations (n=8) at each dilution level.**

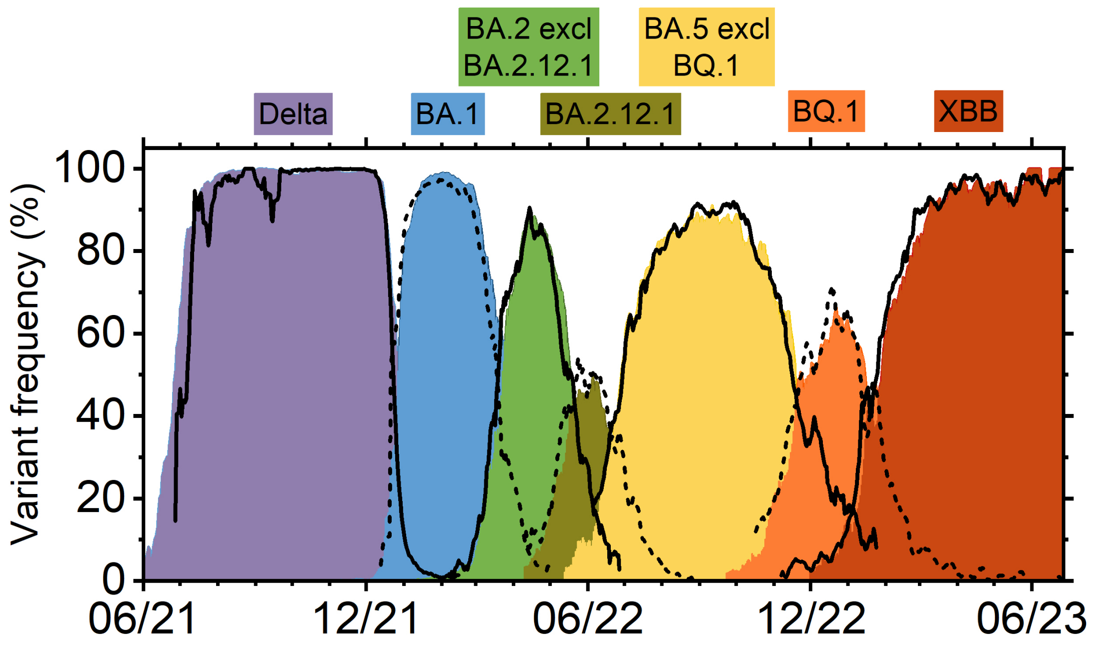


**Figure S2. Variant frequency data with clinical data submitted by "Lab A" from March 21 to April 2, 2022, omitted. The 7-day simple moving average of the frequency of individual variants in clinical sequences collected in Minnesota is shown as consolidated colored bars, vs time. The 7-day simple moving average of the frequency of individual variants in Metro Plant influent is shown in solid or dashed black lines, vs time. Variants are labeled across the top.**

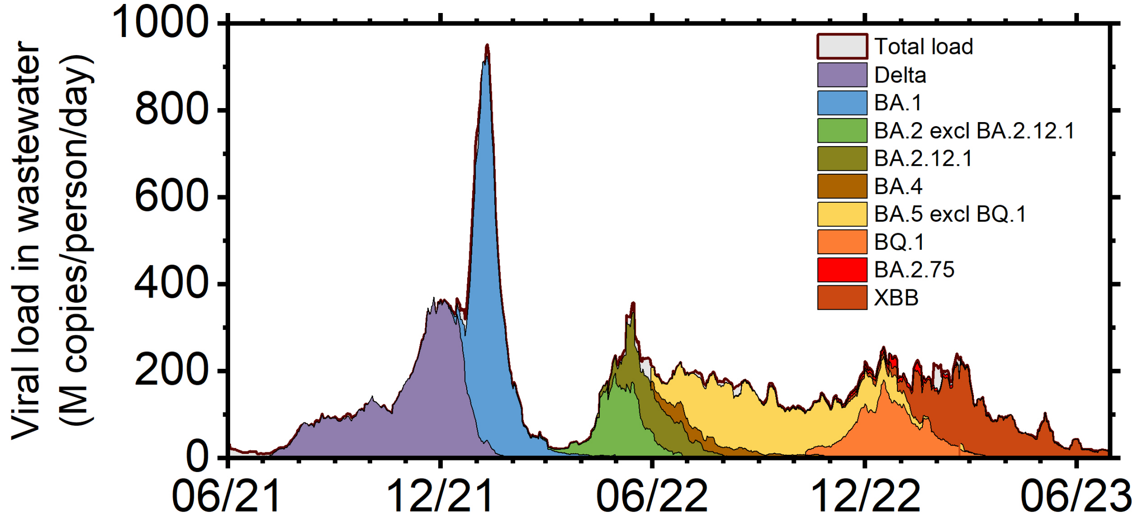


**Figure S3. Stacked area plot of variant loads over time. The 7-day simple moving average of the load of individual variants in Metro Plant influent is shown by the colored areas with black outline, vs time, from June 1, 2021, to June 30, 2023. The 7-day simple moving average of the total daily SARS-CoV-2 load in Metro Plant influent is shown in gray bars with red/black outline, vs. time. The sum of the calculated variant loads over this period comes to 96% of the total load measured using N1 and N2.**
